# User testing to inform modification of the MyHealthyGut digital health application in inflammatory bowel disease

**DOI:** 10.1101/2023.03.23.23287648

**Authors:** Madeline Erlich, Sarah Lindblad, Natasha Haskey, Darlene Higbee Clarkin, Taojie Dong, Ruth Harvie, Genelle Lunken, Jess Pirnack, Kevan Jacobson

## Abstract

**Introduction:** Inflammatory bowel disease (IBD), characterized by chronic intestinal inflammation, can be subcategorized into Crohn’s disease and ulcerative colitis. The treatment for these conditions is unique to each patient, and may include lifestyle changes, pharmaceutical intervention, and surgery. Lifestyle changes, such as dietary intervention, are a cornerstone of IBD symptom management. Given the daily burden of this disease, self-management is paramount in coping with and/or minimizing symptoms. The MyHealthyGut application (app), successfully proven to be a self-management tool for celiac disease, shows promise for use in an IBD patient population.

**Objective:** To undertake user testing to inform the development of an IBD-focused version of the current MyHealthyGut app.

**Methods:** This study was undertaken between October 2021 and April 2022. Participants included IBD patients and healthcare practitioners (HCPs) (physicians, registered dietitians [RD], and registered nurses [RN]), using social media postings and convenience sampling. Two RDs demonstrated how to use the current functions and features of the app with each participant. Participants used the app for a 2-week period which was followed by participation in a focus group or individual interview to provide feedback on the app. Qualitative questionnaires, tailored to each patient category, were administered verbally and feedback was recorded. Thematic analysis techniques were used for data quantification and analysis.

**Results:** 15 participants were recruited and enrolled. Of these, 14 participants took part in the focus group and/or individual interviews. The feedback suggested changes related to clinical uses (e.g. incorporating information collected by the app into electronic medical record systems), food and symptom tracking (e.g. the option to track water intake), ease of use (e.g. the option to autofill food tracker with frequently consumed meals), and app content (e.g. information about IBD treatments). All (100%) of participants reported that they would either use the app themselves or recommend the app to patients, once their suggestions were implemented.

**Conclusion:** Through user testing and feedback collection, priorities for app modification were identified. Areas of modification in the app functions and features, ease of use, and content were identified. Once updated to meet the needs of IBD patients, the MyHealthyGut app may be a useful tool for IBD self-management.

**Funding Source:** Canadian Foundation of Dietetic Research

## Introduction

Inflammatory bowel disease (IBD) is characterized by chronic intestinal inflammation, broadly categorized into two types, Crohn’s disease (CD) and ulcerative colitis (UC). Typical IBD symptoms include diarrhea, abdominal pain, weight loss, and bloody stools. The etiology of IBD has not been fully elucidated however environmental factors, such as diet has been under the spotlight in IBD development and management in recent decades [4,5]. Multiple studies have shown evidence that a diet high in fibre, fish, fruits, and vegetables has a protective effect against IBD. Conversely, a typical western diet, characteristically rich in n-6 polyunsaturated fatty acids (PUFA) and low in fibre, is suggested to contribute to the onset of IBD [6, 7]. Moreover, the rise in IBD incidence in countries adopting westernized practices, such as Africa, Asia, and South America, further supports the potential impact of the western diet [8].

Though IBD is seen globally, the prevalence of IBD was found to be the highest in highly industrialized countries, including those in North America, Australasia, and parts of Europe [8]. In Canada, the prevalence of IBD in 2018 is 0.7% and is forecasted to increase to 1.0% by 2030 [9]. The IBD patient population grows in tandem with its financial burden on the healthcare system, predominantly through prescription drugs, biological therapies, and hospitalizations [10]. These direct healthcare costs, defined as the cost of medically necessary services for each IBD patient, were estimated in 2018 to be $14.6 billion (USD) and $1.29 billiion (CAD) in the US and Canada, respectively [10]. In addition to direct healthcare costs, indirect costs include those related to patient time off work (absenteeism), decreased productivity, the mental health burden of living with a chronic disease, and premature and long-term disability [10]. One solution to the costs of primary care to both IBD patients and the healthcare system is a greater emphasis and investment in self-management strategies.

With the rise of mobile phone users worldwide, the electronic health (eHealth) monitoring movement is at the forefront of disease prevention and self-management. The eHealth applications (app) developed for chronic disease management including diabetes, several types of cancer, and chronic obstructive pulmonary disease, have demonstrated efficacy through decreased hospitalization rates, healthcare costs, and increased patient sense of security, understanding of their condition, and convenience [11]. The established benefits of eHealth in chronic disease management show promise for use of a mobile app as a tool in IBD management. A focus group study conducted by Khan and colleagues (2016) highlighted the IBD patient’s need for a tool with better symptom tracking, disease control assessment, medication adherence, physician feedback, objective setting, and education [12]. While the validation of the IBD management mobile app, HealthPROMISE, created based on Khan and colleagues study outcomes revealed a reduction in hospitalization and increased understanding of IBD, however, no significant change in IBD quality indicators such as bowel symptoms, emotional health, social function, and overall quality of life were seen [13]. Given the importance of diet in IBD management and the lack of scientific-based nutrient advice and analysis features in current IBD management mobile apps, there is a need for a diet-centred mobile app, providing dietary guidance and support to IBD patients.

MyHealthyGut (MHG) is a mobile app that was originally designed for celiac disease (CeD) self-management [14,15]. At present, the only treatment for CeD is a strict gluten-free (GF) diet [16]. The app was developed to provide guidance, diet and symptom tracking, and information to users, related to the GF diet and CeD. Although CeD and IBD have key distinctions (primarily in the way that CeD is gluten-induced immune-mediated enteropathy), they may share similar risk association, etiology and patient experience [17,18,19]. Given these similarities, we hypothesized that many of the functions and features of the original MHG app, such as diet and symptom self-monitoring tools, may be transferable and beneficial to IBD patients, once tailored to the disease. The purpose of this study is to investigate the ways in which the original MHG app can be modified and tailored to meet the needs of IBD patients and HCPs.

## Methods

### Recruitment

This project was completed between October 2021 - April 2022. We recruited participants using a flyer posted on social media and convenience sampling. Participants included IBD patients and HCPs specializing in IBD (i.e., registered dietitians [RD], physicians, and registered nurses [RN]).

### Ethics Approval

The project was reviewed and approved by the Research Ethics Board of University of British Columbia (H21-02662).

### Study design

After completing the consent process, participants met with two RDs at the beginning of the study period. Participants were guided through the app setup, as well as the key functions and features of the app. Participants were instructed to test the app for a two-week period and make note of what they liked or disliked about the app features and functions, as well as any ideas for improvement. Following the app testing period, the study RDs interviewed participants (grouped by type) in either a focus group or individual interview, guided by questionnaires developed for the purposes of this study. Participants received a $25.00 CAD gift card to Amazon, to provide compensation for their participation in the study.

### Questionnaire development

Three versions of an open-response qualitative interview-based questionnaire were developed, specific to participant type (patient, physician, RD/RN), for feedback provision. Common mHealth evaluation questionnaires, including SUS, TAM, and uMARS, were used as resources in the development of our questionnaires [19].

#### Patient version (11 questions)

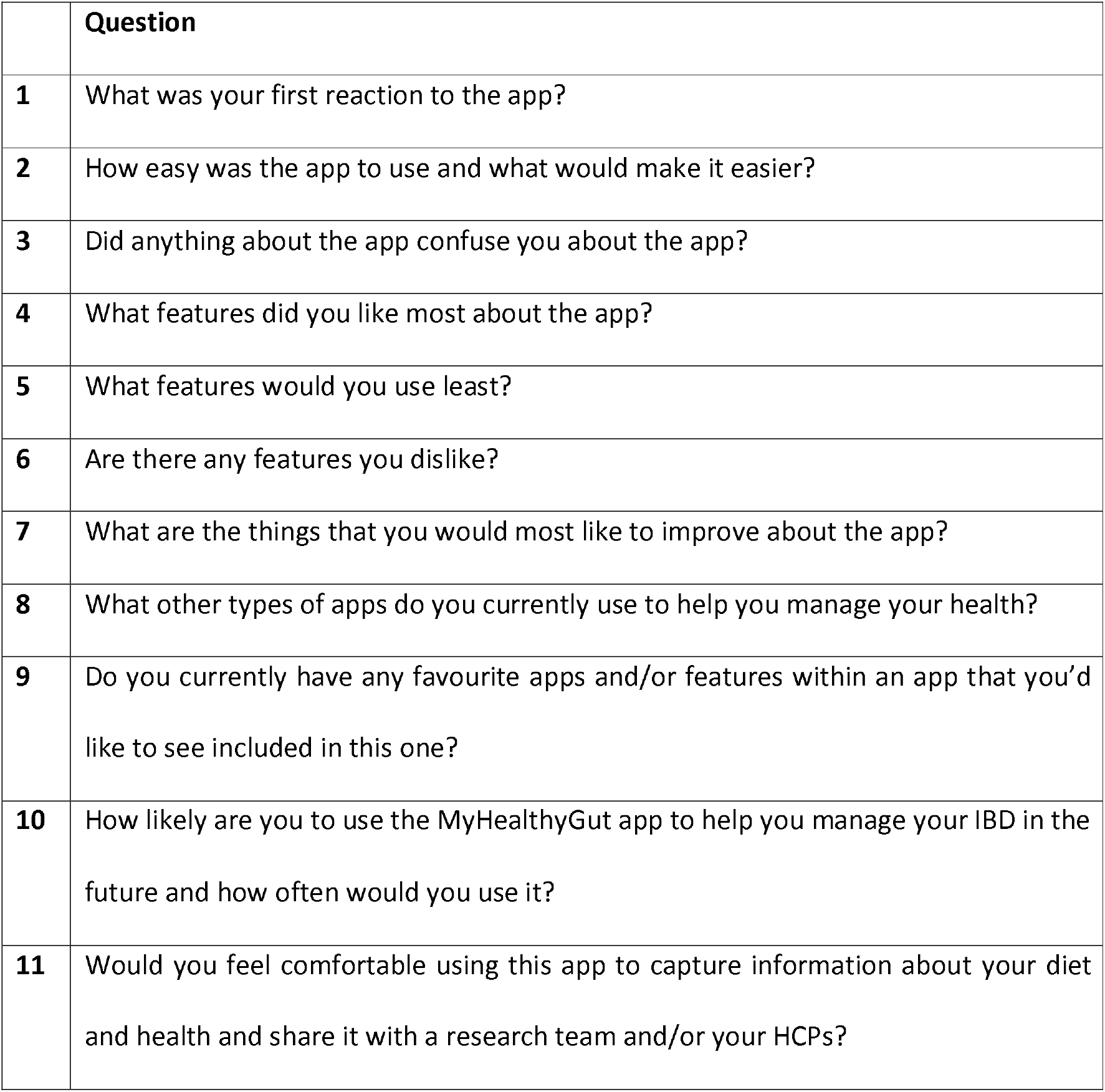

|  | Question |
| --- | --- |
| 1 | What was your first reaction to the app? |
| 2 | How easy was the app to use and what would make it easier? |
| 3 | Did anything about the app confuse you about the app? |
| 4 | What features did you like most about the app? |
| 5 | What features would you use least? |
| 6 | Are there any features you dislike? |
| 7 | What are the things that you would most like to improve about the app? |
| 8 | What other types of apps do you currently use to help you manage your health? |
| 9 | Do you currently have any favourite apps and/or features within an app that you'd like to see included in this one? |
| 10 | How likely are you to use the MyHealthyGut app to help you manage your IBD in the future and how often would you use it? |
| 11 | Would you feel comfortable using this app to capture information about your diet and health and share it with a research team and/or your HCPs? |

#### Physician version (9 questions)

Physician version (9 questions):
|  | Question |
| --- | --- |
| 1 | How often do you see your average patient? |
| 2 | Do you have a series of questions you ask your patients in order to understand their disease state? |
| 3 | How beneficial are disease activity indices in your practice and would it be beneficial if an app collected all components of that algorithm regularly to produce a score? |
| 4 | What are the key areas of concern for your patients when it comes to managing their IBD? |
| 5 | What are some functionalities an app would need in order to be beneficial to your practice? |
| 6 | Would it be valuable for the app to flag a pattern or the frequency of specific symptoms and suggest the patient seek medical attention? |
| 7 | Are there any new innovations related to self-management of IBD that you would like to see in an app like this? |
| 8 | Do you recommend any apps to your patients or their families, related to IBD self-management? |
| 9 | Are there any specific resources you recommend to your patients or their families, and if so, which ones? |

#### RD/RN version (9 questions)

RD/RN version (9 questions):
|  | Question |
| --- | --- |
| 1 | What are the key areas of concern for your patients when it comes to managing their IBD? |
| 2 | Do you recommend any apps to your patients or their families, related to IBD self-management? |
| 3 | What features of the MyHealthyGut app did you like most and think your patients would use most? |
| 4 | Are there any features you disliked about the app, or think your patients would use the least? |
| 5 | Any improvements that you would suggest for the symptom tracker? (6) What kind of educational content would you like to see within the app? |
| 6 | What kind of educational content would you like to see within the app? |
| 7 | Are there any features you think would be important for your patients to have within an app for IBD self-management? |
| 8 | Would you recommend this app if it were tailored to IBD self-management? |
| 9 | Would you find it useful if patients could share information about their diet and health through the app ahead of appointments? |

### Data Analysis

Conversations were recorded and transcribed by the interviewers. Thematic analyses were performed in accordance with the Kiger and Varpio (2020) “Thematic analysis of qualitative data: AMEE Guide No. 131”, and was completed as follows:

Step (1) *Familiarize yourself with the data*: The RDs reviewed the interview transcripts.

Step (2) *Generating initial codes*: Codes were created and defined to categorize the content of each transcript.

Step (3) *Searching for themes*: Through completing steps 1 and 2, themes were identified independently.

Step (4) *Reviewing themes*: The RDs reviewed the proposed themes together.

Step (5) *Defining and naming themes*: The identified themes were named and defined.

Step (6) *Producing the report/manuscript*: A report was created based on the analysis of the frequency of each theme arising in the discussion and specific ideas or pieces of feedback within each theme.

## Results

### Participants

A total of 15 participants (9 patients, 6 HCPs, including 2 MDs, 3 RDs, and 1 RN) were recruited to participate. 14 participants provided feedback, however, 1 patient participant’s feedback was excluded, as it was related to the participant’s person use of supplements and was therefore unrelated to the objective of the study.

### Themes

Feedback collected was grouped into 12 different themes, based on the type of comments and answeres received: 1) clinical use, 2) symptom management, 3) nutrition, 4) medications/supplements, 5) parents/transitional phase, 6) customization, 7) resources/education, 8) functionality, 9) accessibility, 10) community, 11) wellness, and 12) research. Table 1 summarizes the frequency of theme occurrence. In summary, the MDs provided feedback most frequently about the clinical use theme. The RDs provided feedback most frequently about the nutrition, symptom management, and resources/education themes. The RN provided feedback most frequently about the nutrition and resources/education themes. The patients provided feedback most frequently about nutrition, symptom management, resources/education, and the functionality/ease of use themes.

**Table 1.** Frequency of Theme Occurrence by Participant Type

| **Themes** | **Physician**<br>**(n=1)** | **RDs**<br>**(n=3)** | **RN**<br>**(n=1)** | **Patients**<br>**(n=8)** |
| --- | --- | --- | --- | --- |
| Clinical use | 14 | 9 | 2 | 12 |
| Symptom Management | 5 | 23 | 4 | 26 |
| Nutrition | 2 | 31 | 12 | 29 |
| Medications/Supplements | 0 | 14 | 4 | 3 |
| Parents/Transitional Phase | 0 | 0 | 3 | 1 |
| Customization | 0 | 7 | 5 | 14 |
| Resources/Education | 0 | 27 | 17 | 24 |
| Functionality | 0 | 12 | 6 | 29 |
| Accessibility | 0 | 10 | 0 | 6 |
| Community | 0 | 3 | 0 | 8 |
| Wellness | 0 | 12 | 0 | 7 |
| Research | 0 | 1 | 0 | 6 |

The 12 themes were defined as follows:

1. *Clinical Use*: How a HCPs could use the app in their practice or to connect with their patients/other HCPs
2. *Symptom Management*: Any feedback or commentary related to symptoms, the symptom tracker, and/or symptom management
3. *Nutrition*: Any feedback or commentary related to nutrition for IBD and/or the food tracker
4. *Medications and Supplements*: Any feedback or commentary related to the use of medication or supplements for the prevention and treatment of symptoms of IBD and other conditions
5. *Parents/Transitional Phase*: Any feedback or commentary related to a parent using the app on behalf of their child with IBD
6. *Customization*: Any feedback or commentary related to the ability of each user to customize the app to meet their unique needs
7. *Resources/Education*: Any feedback or commentary related to topics of interest that could be included in the app as a source of information
8. *Functionality*: Any feedback or commentary related to the app ease of use
9. *Accessibility*: Any feedback or commentary related to barriers of accessibility to the app
10. *Community*: Any feedback or commentary related to related to ways in which users could connect with others on the app
11. *Wellness*: Any feedback or commentary related to wellness outside of ‘clinical’ IBD health (i.e., mental wellness, physical activity)
12. *Research*: Any feedback or commentary related to access to future research studies, new advances in research, etc. through the app

### Representative constructive feedback quotations for app modification

1. “Definitely the meal and symptom tracker is what I geared most towards. I think if there was a water tracking piece to it, that would’ve been better for me.” - IBD patient
2. “I didn’t really use ‘Ask a Question’, but that’s the only feature I didn’t use.” - IBD patient
3. “[If the app had everything I wanted], the goal would be to use it once a day.” - IBD patient
4. “Breaking the food tracker down into times, rather than just meals [would improve useability].” - IBD patient
5. “When I search for food [in the tracker], that’s where I can imagine your typical day already being there. And then it could ask ‘Was it a typical day?’ [and you’d respond] ‘yes’ or ‘no’.” - IBD patient
6. “I can see the safe food list being beneficial to our patients. Gluten is a problem for some, but more so FODMAP and fibre content. Tagging insoluble vs. soluble fibre in the list would be a super good resource.” - RD
7. “As far as educational content, we talk a lot with patients about managing symptoms. Things that as dietitians we can take for granted, such as eating small, frequent meals.” - RD
8. “It would be best if the results [from the app trackers] could be integrated into our Electronic Medical Records” - Physician
9. “I would definitely [recommend the app to my patients if it were tailored to IBD]. It’s nice to have a one stop shop.” - RD
10. “I would never delete the app if it had the function of communicating distressing symptoms to my gastroenterologist” - IBD patient

### Application usability

All participants (n=14) said that they would be eager to use and/or recommend an IBD-focused self-management app if it came to fruition and reflected their suggestions.

## Discussion

### Principal Results (Priorities)

The most frequently mentioned theme sub-topics were identified and summarized, with the intention of providing guidance for prioritization in the MHG app modification phase that will follow on from this study. Identified priorities were divided into two groups: technical priorities and content development priorities. Technical priorities included automation, customization, food tracker improvements, symptom tracker improvements, and tracking medications. Content development priorities included the development of various resources and education, improving the language in the app, and creating recipes specific to common IBD trigger foods.

### Strengths/Limitations

A notable strength of this study was the enrolment of participants who either had IBD or worked as a HCP in the IBD field. In doing this, we were able to collect feedback that was very specific to the goal end user of the app. We were also able to capture two different perspectives, the very specific perspective of the patient and the more general and widely applicable perspective of the practitioner. This strategy will allow us to develop the IBD version of the app with user-centred design.

A limitation of the study is the small sample size (n=15). Recruitment was completed only through social media posting and word of mouth, due to financial limitations. With greater funding allocated to recruitment, more resources could have been put towards the strategy used, which could have elicited a larger sample size. Unfortunately, the limited sample in this study affects the generalizability of the results. Another notable limitation is the use of a CeD based app in the trial with an IBD population. When trialling the app, some of the features and functions were irrelevant to the study population. Therefore, participants’ experience of the app was not as thorough as it would have been with IBD-tailored features. Finally, the study was undertaken during the COVID-19 pandemic, which impacted almost all aspects of everyday life, on a global scale. This may have affected recruitment, as well as the level of participant dedication.

### Comparison with prior work

To our knowledge, the is the first study to user test a mobile app for modification as a nutrition therapy app for IBD patients. There are several eHealth platforms focusing on IBD patient education, diagnostic support, symptom management for clinical purpose, yet the outcomes for each study reveal inconsistent effects on IBD quality indicators [19, 21]. The MyHealthyGut app diet tracking feature along with the IBD symptom diary provides users with personalized diet associated patterns linking their food intake and symptoms, which provides an opportunity for more targeted treatment and better treatment efficacy. A study by Villinger and colleagues (2019) demonstrated the poor long-term user adhesion to eHealth platforms, impacting prolonged beneficial outcomes [22]. App usability has been identified as the barrier to long-term user engagement of nutrition app and food database tools in previous studies [23]. In this study, feedback on the usability of the app was assessed through questionnaires and priority changes for a more user-friendly design were identified.

Finally, previous eHealth platforms designed for IBD management have been assessed by IBD patients’ satisfaction and healthcare activities in isolation, which excludes the view of the healthcare team [18, 23]. In this project, the HCPs in the IBD field were included as part of the assessment process and were also able to provide feedback for further app development.

### Future Directions

Our future directions are to implement the identified priority changes to the existing MHG app. Once the new IBD version of MHG exists, the functions and features could be tested by users with similar methodology to the previously completed validation study [14]. We also intend to validate the food and symptom trackers so that they can be utilized in scientific research. Finally, there is potential for future variations of the app to exist for a multitude of other disease states.

## Conclusion

With modifications to functionality and capabilities of the app, as well as the focus of the educational content, the current app is transferable to an IBD population. Responses from focus groups and interviews will be used to inform future modifications in the app. The proposed IBD self-management app would be of value to, and used or recommended by, both IBD patients and HCPs specializing in IBD.

## Data Availability

All data produced in the present study are available upon reasonable request to the authors

## Abbreviations

(IBD): Inflammatory Bowel Disease
(HCP): Health Care Practitioner
(RD): Registered Dietitian
(RN): Registered Nurse
(CD): Crohn’s Disease
(UC): Ulcerative Colitis
(eHealth): electronic health
(MHG): MyHealthyGut
(CeD): Celiac Disease
(GF): gluten free
(app): application

## Acknowledgements

We would like to take the opportunity to thank the patients and health care professionals that provided feedback on the MyHealthyGut app.

